# A TRANSPARENT, OPEN-SOURCE SIRD MODEL FOR COVID19 DEATH PROJECTIONS IN INDIA

**DOI:** 10.1101/2020.06.02.20119917

**Authors:** Ananye Agarwal, Utkarsh Tyagi

## Abstract

As India emerges from the lockdown with ever higher COVID19 case counts and a mounting death toll, reliable projections of case numbers and deaths counts are critical in informing policy decisions. We examine various existing models and their shortcomings. Given the amount of uncertainty surrounding the disease we choose a simple SIRD model with minimal assumptions enabling us to make robust predictions. We employ publicly available mobility data from Google to estimate social distancing covariates which influence how fast the disease spreads. We further present a novel method for estimating the uncertainty in our predictions based on first principles. To demonstrate, we fit our model to three regions (Spain, Italy, NYC) where the peak has passed and obtain predictions for the Indian states of Delhi and Maharashtra where the peak is desperately awaited.

## 1 Introduction

India has just emerged from a long and strict lockdown. There are doubts about effective the lockdown has been and where the country is going from here. Given the steep economic cost of lockdowns it is important to understand their impact on case numbers and death counts. Further, given the lack of information about the novel coronavirus it is extremely hard to model the disease realistically. In our work, we choose a simple SIRD compartmental model in order to model death counts due to COVID19.

At a high level, the SIRD compartmental model in epidemiology partitions the population into four compartments susceptible (those people who can potentially be infected from the virus), infected, recovered and dead. It also takes as input the the Reproduction Rate *(R*_0_) which depends on the social distancing behaviour of the people in the country, This allows us to vary for the infection by capturing details of the policies being implemented with respect to lockdowns in the country and summarizing it in a single number.

**Figure 1:**
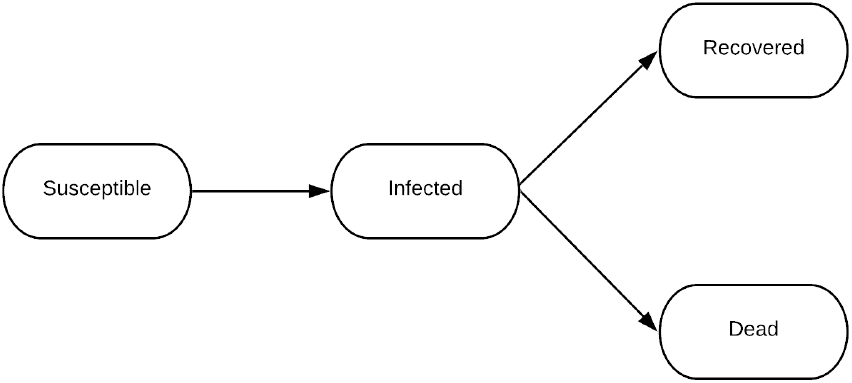
Block Diagram for SIRD Model

## 2 Related Work

We list down popular models being referred to by the Indian and international community and their shortomings, and then we discuss how we overcome them.

### IHME Model

After getting cited by the United States Government for the COVID19 projections, the IHME model [1] has received a lot of attention from the public and professional community. It relies on data from China and Italy for training the model parameters and extend the model to the United States. One of the criticisms for the model is that it uses statistical definitions to make predictions. It cannot give information on the actual parameters underlying the disease spread due to departure from epidemiological theory. It has consistently under-performed compared to the actual statistics, and needs to be re-updated after every few weeks. The code for this model is only partly open-sourced, it is very hard for a third-party to replicate their results.

### Multi-Compartment Model

Overcoming the limitations of the statistical models like IHME Projections, Stanford published a nine compartment model [2], which explicitly tracks nine compartments, including exposed, asymptomatic, pre-symptomatic, symptomatic, hospitalized, and recovered. A similar model was developed in the Indian context. [3] The criticism of this complex epidemiological model is that they have a large number of tunable and learnable parameters which interact in unpredictable ways. Further, given the uncertainty surrounding the disease it is not clear how these values can be fixed reliably. Since the data provided by the government becomes unreliable as the cases increase, the modelling in such cases becomes unreliable.

### PRACRITI Model by M3RG, IIT Delhi

The model by M3RG Group of IIT Delhi [4] is an extension of the SIER Model that creates Adaptive, Interacting and Cluster-based, SEIR compartments for district level populations. This model suffers from limitations as stated for both previous models. They have added a lot of mathematical parameters with do not have physical meaning, and thus cannot be checked against real-world data. With an additional migration term, they assume the migration statistics irrespective of the movement of the migrants and their social distancing measures. Next, the model doesn’t perform well as on simple inspection. As of the time of writing, the *R*_0_ for Mumbai is reported as 0.73, while same value for the State of Maharashtra is 1.33. This is unreasonable because Mumbai is known to be contributing the most to the case load of Maharashtra. Further, there are no confidence intervals in their plots which makes their predictions meaningless since it is impossible to extrapolate cases with 100% accuracy. Further, they have not open-sourced their code.

## 3 Novelty

Our model is inspired by the ones mentioned above and aims to combine various techniques to avoid the pitfalls outlined above. Salient features of our model include -

### Modelling on Daily Deceased Data

We use daily deceased data for time-series forecasting of the COVID19. In this work, we only include projections for future deaths, although this can be extended to projections for other things like the number of infections and the number of ICU beds required. The benefits of using deaths rather than infections is that the latter crucially depends on the scale of testing and reporting by the relevant government agencies. Further, cases are reported as and when they are discovered which means that the time when a person was infected does not show up in the data. On the other hand, deaths due to COVID19 occur in hospital ICUs and the exact time of death is known. Further, since all patients who eventually die end up in the hospitals it is unlikely that deaths are being under-counted (barring deliberate under-reporting). In this way, we bypass the uncertainty of testing. Further, it can be argued that the death toll and number of ICU beds required is more important to estimate than the total number of infections since most infected people recover from COVID19 without medical assistance.

### Simplicity

The model implements a four compartment SIRD based model where we vary reproduction number (*R*_0_) with respect to the social distancing measures. This means that *R*_0_ effectively summarizes the entire suite of prevention strategies adopted by a region as well as migration and mixing patters. This gives us flexibility to monitor real-time policy changes in the data and update *R*_0_ accordingly. Also, because of just 4 compartments we require only a few disease-specific parameters (the infectious period and mortality rate). More complicated models need a larger number of parameters. Since the values of these parameters are not known accurately at this point of time, it makes these models prone to over-fitting. A fewer number of parameters also means that our model is region agnostic and can be implemented on national and state levels, all over the world with minimal modification.

### Transparency

Since our model is based parameters well-documented in epidemiological theory, we can do a sanity check on the inferred values to see if they agree with what is known at this point of time. This can also be used in principle to compare the effects of different policies in mitigating the spread of the virus by comparing the variation in the reproduction number. Further, we believe that any modelling effort must strive to be as transparent as possible. This is because to the non-expert, the projections churned out by sophisticated mathematical machinery seem to carry more weight than they really do. In reality, given the large number of variables involved, most mathematical models end up being nothing more than educated guesses and are only as strong as the assumptions they implicitly make. Therefore, we believe it is critical that all results should be made publicly available and the methodology explained in as detailed a manner as possible. Many models we have mentioned above do not do this. They have not open-sourced their code completely and it is not possible, or is very difficult and time-consuming to replicate their results by reading the technical reports alone. In contrast, we have completely open-sourced our code with relevant documentation so that the community can critique our assumptions and contribute their own ideas to improve the model.

### Explicit CIs

Since it is impossible to predict with 100% accuracy the death count into the future, no model can be complete without providing confidence intervals for its projections. We present a novel strategy to compute these confidence intervals from first principles with the empirical claim that *most* of the time the observed death counts will fall within these intervals.

## 4 Description of the Model

### 4.1 Mathematical formulation

An SIRD model is described by the following coupled differential equations,

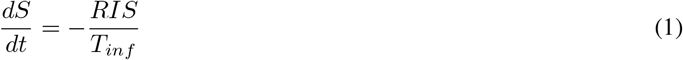

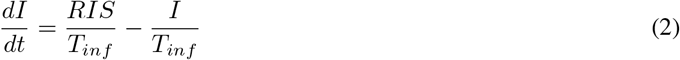

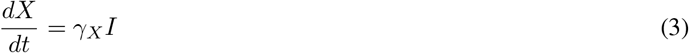

Here, *S*, *I*, *X* are respectively the fraction of the population that is susceptible, infected and deceased. We omit the number of people who have recovered because we do not fit our model on that data.

*T_inf_* denotes the median amount of time a person stays infectious, *γ_X_* is average number of people who die from COVID19 in a day as a fraction of the total number of active cases on that day. *R* is the reproduction number of the disease which measures the average number of people an infected person transmits the disease to. *R* > 1 implies that the case count rises over time while *R* < 1 implies that the case count diminishes over time with the rate of spread being determined by *R*. Note that in general *R* varies with time depending on the extent of social distancing practiced.

Note that *γ_X_* and *T_inf_* are spatially-invariant and time-invariant properties of the disease. They depend on the virus specifics and how the human body responds to it. Thus, this parameters can be bounded in a small range based on preliminary studies from Wuhan and Europe, where cases of COVID19 have been large. Studies by Wilson [5] on the data from New York city shows that the Infection Fatality Rate (IFR) for United States is 0:863%. Similar estimate done for the COVID19 outbreak on Diamond Princess cruise ship [6] reveals that IFR was 1:2%(0:38% 2:7%). Next, the study by Bar-On et al. [7] shows that even if recovery time for the infection is 2-3 weeks, the infectious phase for an individual is 4-5 days on average. We use these bounds for *γ_X_* and *T_inf_* in the grid search. Empirically, the model performs best for IFR and *T_inf_* as 0:8% and 5 days respectively.

We then see that all the information about the progression of the disease lies in a single parameter - *R*. This is the major advantage of using a simple model. We do not have to deal with lots of uncertain parameters that influence the final curve in unpredictable ways, we can focus on estimating *R* as best we can. Further, *R* is a well-established measure of disease spread in epidemiological literature which means there are already many existing estimates of *R* which our model can leverage.

### 4.2 The parameter *R*

*R* is a function of time and depends on (among other things), the lockdown and social distancing measures adopted by each country. To estimate *R* we leverage open-source, real-time social distancing data published by Google [8], which allows us to model various mitigation measures by just two parameters as described below. While the social mobility data does not account directly for various measures such as contact tracing and mask usage, we nonetheless postulate that the timing of these measures is correlated with the timing of social distancing measures indicated by the mobility data.

The data, available in aggregated form, shows how the number of visitors who go to (or spend time in) categorized places change compared to pre-COVID days. A baseline day represents a normal value for that day of the week. The baseline day is the median value from the 5-week period Jan 3 – Feb 6, 2020. The places are categorized into Retail and Creation, Grocery and Pharmacy, Parks, Transit Stations, Workplaces, and Residential.

**Table 1:**
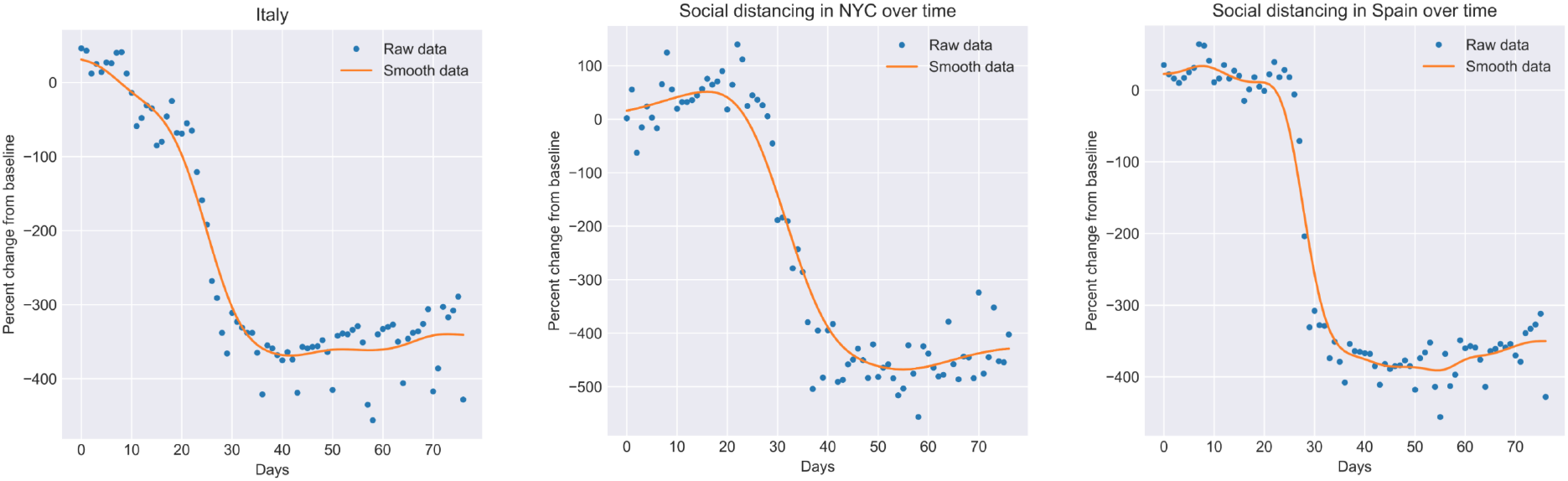
Raw and smooth social distancing data for three different regions from 15 Feb

Additionally, for a sanity check, we looked at the smartphone penetration in the country to validate the model. The report by StatCounter [9] suggests that Android based smartphones constitute more than 95% of all smartphones being used in the country in April 2020. With this size of market share, the open source data-model performs well.

We construct a social distancing covariate *s*(*t*) from the changes in social mobility at various locations

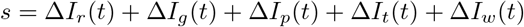

where each of the terms on the RHS above denote percentage change from baseline in mobility at the following locations

- Δ*I_r_* (*t*) - retail and recreation
- Δ*I_g_* (*t*) - grocery and pharmacy
- Δ*I_p_* (*t*) - parks
- Δ*I_t_* (*t*)-transit stations
- Δ*I_W_* (*t*) - workplaces

Note that we ignore residential mobility data as residential mobility does not contribute to the spread of the disease. Next, we smoothen the covariate *s*(*t*) by applying a Savitzky-Golay filter followed by convolution with a localized Gaussian multiple times. The goal is to smooth out weekly variations in the data but not distort the overall profile of the curve. This gives us 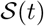, the smooth social distancing covariate.

Since we only care about the timing of social distancing measures, to relate 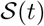 to *R* we introduce two parameters *R_min_*, *R_max_*, the *R* values when 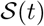 is minimum and maximum respectively. We then define *R* as a linear interpolation function of 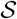 between these two values. Mathematically,

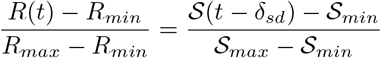

where 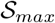, 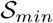 are the global maximum and minimum values of 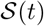. Further, we introduce a fixed lag *δ_sd_* which equals the median time from infection to death. This is because 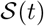 influences the number of infections at time *t*, the effect on deaths is seen only later. Where social distancing data is not available, we naively extrapolate the existing data into the future as well as the past. Concretely, we assume that past values equal the earliest value we know and future values equal the latest value. This amounts to assuming that existing social distancing measures will continue into the future. This assumption can be altered as we learn more about the disease and mitigation strategies in the future.

#### 4.2.1 Fitting the model

We solve the differential equations using a simple iterative procedure where the values of the next day are determined by the values of the previous day.

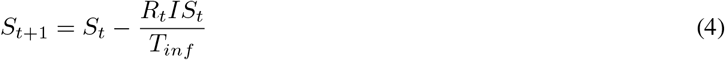

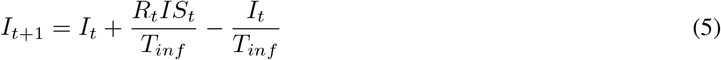

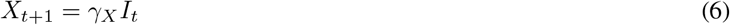

We avoid more complicated numerical techniques like the Runge-Kutta methods because they proved to be too computationally intensive to fit a large number of models. Additionally, for our purposes the above recurrences yield a reasonable approximation.

Note that to solve the system of differential equations above we need to specify an initial condition. In particular we need to specify initial values for time *t*, and each of *S*, *I*, *X*. Since the set of differential equations (1) - (3) is valid at all points of time we can arbitrarily choose a starting point.

We start the model just before we get the first death. Obviously, 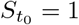, 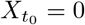. We choose 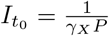 where *P* is the population. This choice implies that at day 1, there will be exactly one death. Since real death counts are discrete, we choose *t*_0_ in a narrow interval around where the actual death count start to rise.

To prepare the daily death counts, we obtain raw death counts from two sources - John Hopkins [10] and the covid19india.org [11], a volunteer-driven tracker project. We then smooth this death count using a combination of Savitzky-Golay and Gaussian convolution filters. Care needs to be taken to not distort the peak too much as with a large amount of smoothing the peak tends to decrease in height.

Finally, to fit the data we do a fine-grained brute force grid search [12] over the possible parameter values we provide and obtain a prediction with the lowest mean squared loss. In general, we fix *γ_X_* = 1.6e–3 (this implies a mortality rate of 0.8%), *δ_sd_* = 23, vary *t*_0_ within a small margin near the beginning of the death count curve and vary *R_max_* from 1.4 – 2.8 and *R_min_* from 0.7 – 0.95.

Predicting the peak (height and position) are quite tricky because the beginning of the curve looks quite similar for different values of *R*. Further, the peak can be quite sensitive to the values of *R_max_* and *R_min_*. *R_min_* is especially hard to estimate because it depends on the death count close to or after the peak, after social distancing measures have been put into place. We discuss these issues in greater detail in the following section.

### 4.3 Uncertainty Analysis

Let M be the random vector corresponding to the choices for (variable) parameters in the model. Then, the density function ƒ(m) constitutes a prior on the choice of these parameters. As an approximation, we assume uniform priors on the parameters (this can in principle be extended to other priors). This is reasonable because from our knowledge of other countries, we can place bound *R* quite confidently whereas pinpointing a single value of *R* is very hard.

Further, we assume that the data 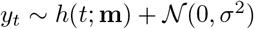, where *h* is the hypothesis (SIRD model), *y_t_* is the observed deaths on day *t* and y is the vector of observed deaths. Note that

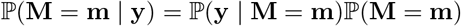

We can assert that we only include parameters in our confidence interval which have probability atleast ∊

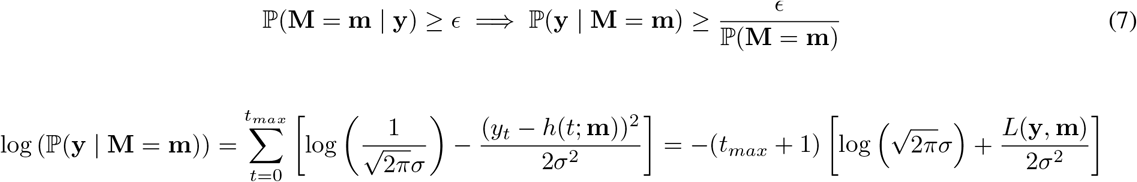

where *L*(y, m) is the average root mean squared loss between y, m. This means that (7) is equivalent to

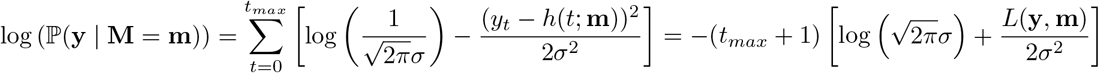

Simplifying this we get,

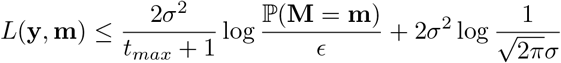

Note that because of the uniform prior the RHS is independent of m. Further, for a theoretical perfect fit, ∊ = 1 and ℙ(M = m) = 1, making the first term zero. Therefore, we can interpret the second term 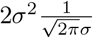 as the minimum loss (or the loss of the best fit curve). This gives us a metric for choosing admissible values of the parameter m.

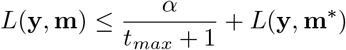

where a is a constant we choose and m* are the best fit parameters. Since our brute-force algorithm gives us the average loss for each possible m, we can select those m for which the loss satisfies the above inequality.

Having obtained a set of values of m, we obtain the corresponding curves for them and plot the minimum and maximum predictions for all these curves to obtain a confidence interval. Note that the acceptable range of *L*(y, m) grows smaller as *t_max_* increases i.e. as we get more data. This agrees with our intuition, which says that as we get more data the confidence interval should become narrower (for fixed *τ*, *α*)

In practice, we start with a conservatively high value of *α* (=200). As we get more data, we increase *α* if the actual values fall outside the confidence interval. The initial value of *α* is chosen based on empirically fitting the model to different countries and observing that this value gives reasonably sized confidence intervals.

## 5 Results

Here we present results for three different regions whose death count peaks have passed. All three - Spain, NYC, Italy were badly effected by coronavirus as India is likely to be. We use these curves to validate the values we have chosen for the fixed parameters and demonstrate the effectiveness of our model. More detailed plots can be found on the Git repository.

Table 2 contains the graph for our projections. Each row contains projections for one region. Across a row, we vary the number data points we fit the model on, and obtain projections for the remaining times and compare them to the actual death counts. Note that in each figure, the area shaded red contains points the model has not been fitted on. The tables 3, 4, 5 contain the numerical values for parameters that are inferred/used by our in each case. Here, train loss is the mean squared loss of the solid blue line with respect to the data it is fitted on. Breakpoint denotes the number of data points from the beginning which are included in the train set. For example, a breakpoint of 60 implies that the first 60 datapoints are used for fitting and the rest are ignored.

It is worth noting that for NYC, Italy and Spain our model predicts *R_min_* < 1 indicating that social distancing measures have been effective in these places which is indeed the case. Also notice that as we expect, with more data the uncertainty interval narrows and converges to the observed data.

### 5.1 Predictions

We now include predictions for two critical regions in India - Delhi and Maharashtra, both badly affected by the virus.

Note that the uncertainty intervals for Maharashtra in the beginning of the curve are very high. This indicates that the model does not fit well to the initial part of the curve. This might be a consequence of (a) the fact that Maharashtra is a large state and different parts of the state are affected differently by the virus (a model fit to Mumbai would perform better) (b) The timing of the drop in 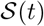 does not track well the timing of prevention measures taken in the state.

On the other hand, the model performs quite well on Delhi. This is likely because Delhi is much more homogeneous in terms of demographics and is more well-connected. This means that Google mobility data is likely to reflect well how much people are social distancing. Note that the current projections in Delhi assume that social distancing will continue at lock-down levels. This is likely to not be the case as Delhi has started re-opening. Nevertheless, the government is still attempting to aggressively identify and quarantine so-called containment zones.

**Table 2:**
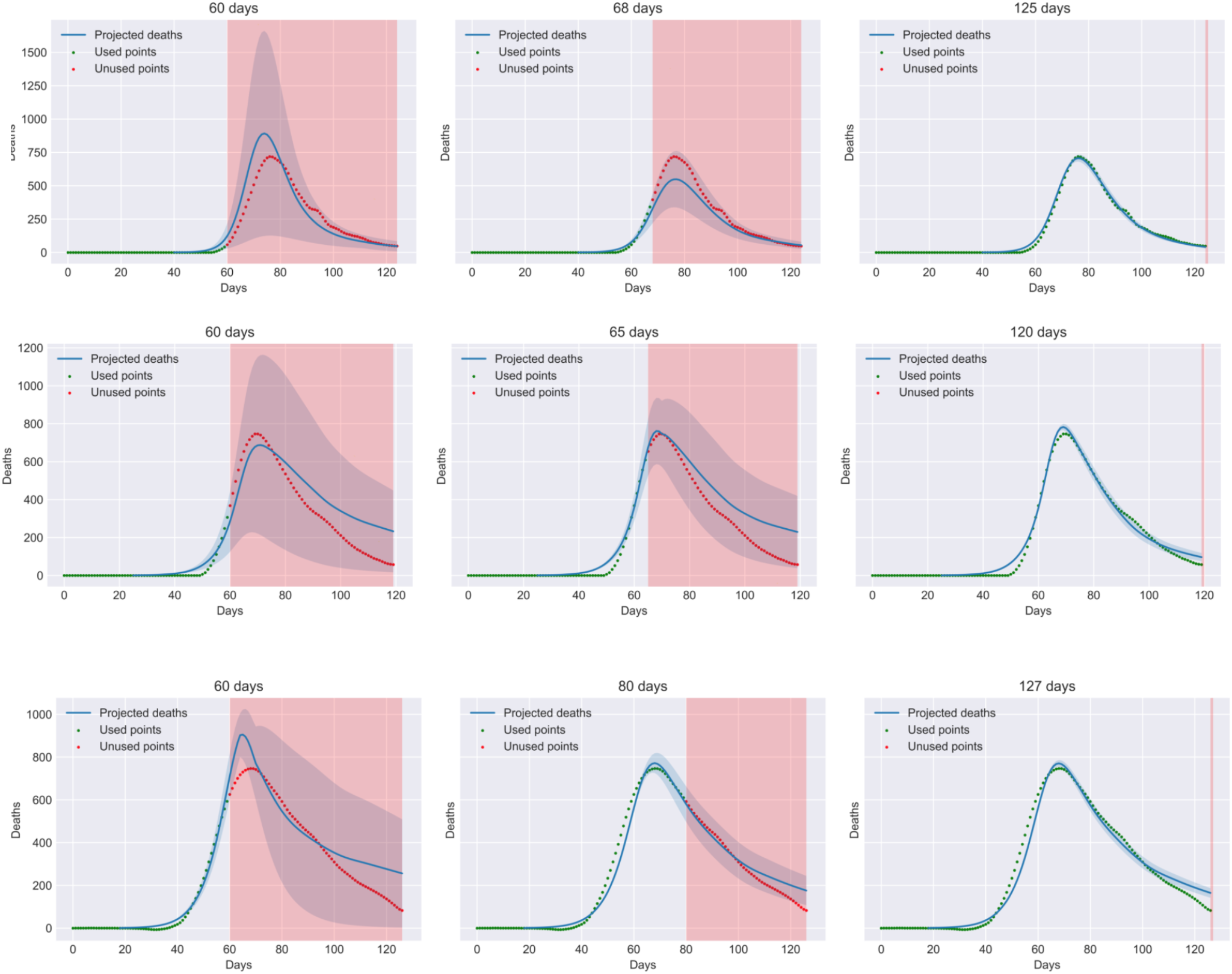
Predictions with uncertainty intervals for three different regions New York City (NYC), Spain, and Italy (from top to bottom). In each figure, the red area contains points the model has not been fitted on and the shaded blue region is a confidence interval.

**Table 3:**
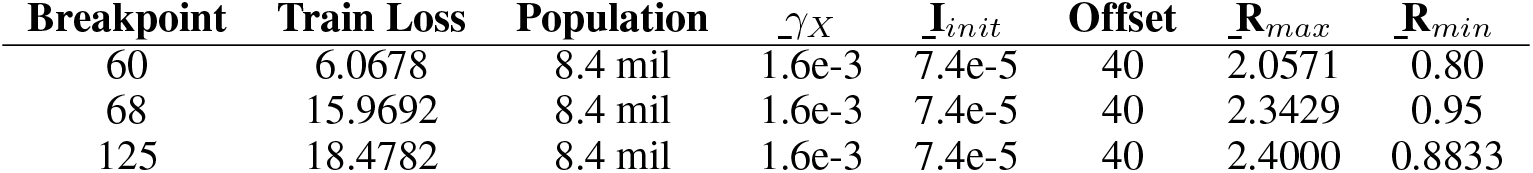
Model Parameters for NYC

**Table 4:**
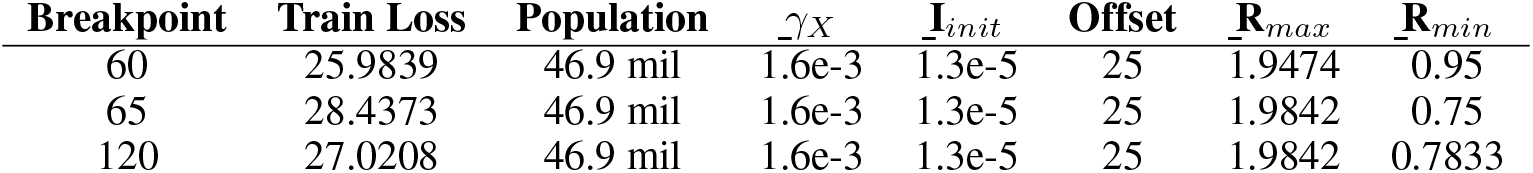
Model Parameters for Spain

## 6 Conclusion

A clear conclusion from the data is that even during the lock-down which has been called one of the strictest in the world, *R_min_* remained above 1, unlike in other countries. This can indeed be seen from our model as well which predicts

**Table 5:** Model Parameters for Italy

| Breakpoint | Train Loss | Population | $\gamma_X$ | $I_{init}$ | Offset | $R_{max}$ | $R_{min}$ |
| --- | --- | --- | --- | --- | --- | --- | --- |
| 60 | 22.9539 | 60.4 mil | 1.6e-3 | 1.1e-5 | 18 | 1.9895 | 0.5 |
| 80 | 41.9706 | 60.4 mil | 1.6e-3 | 1.1e-5 | 18 | 1.9158 | 0.8868 |
| 127 | 40.9382 | 60.4 mil | 1.6e-3 | 1.1e-5 | 18 | 1.9158 | 0.8611 |

**Table 6:**
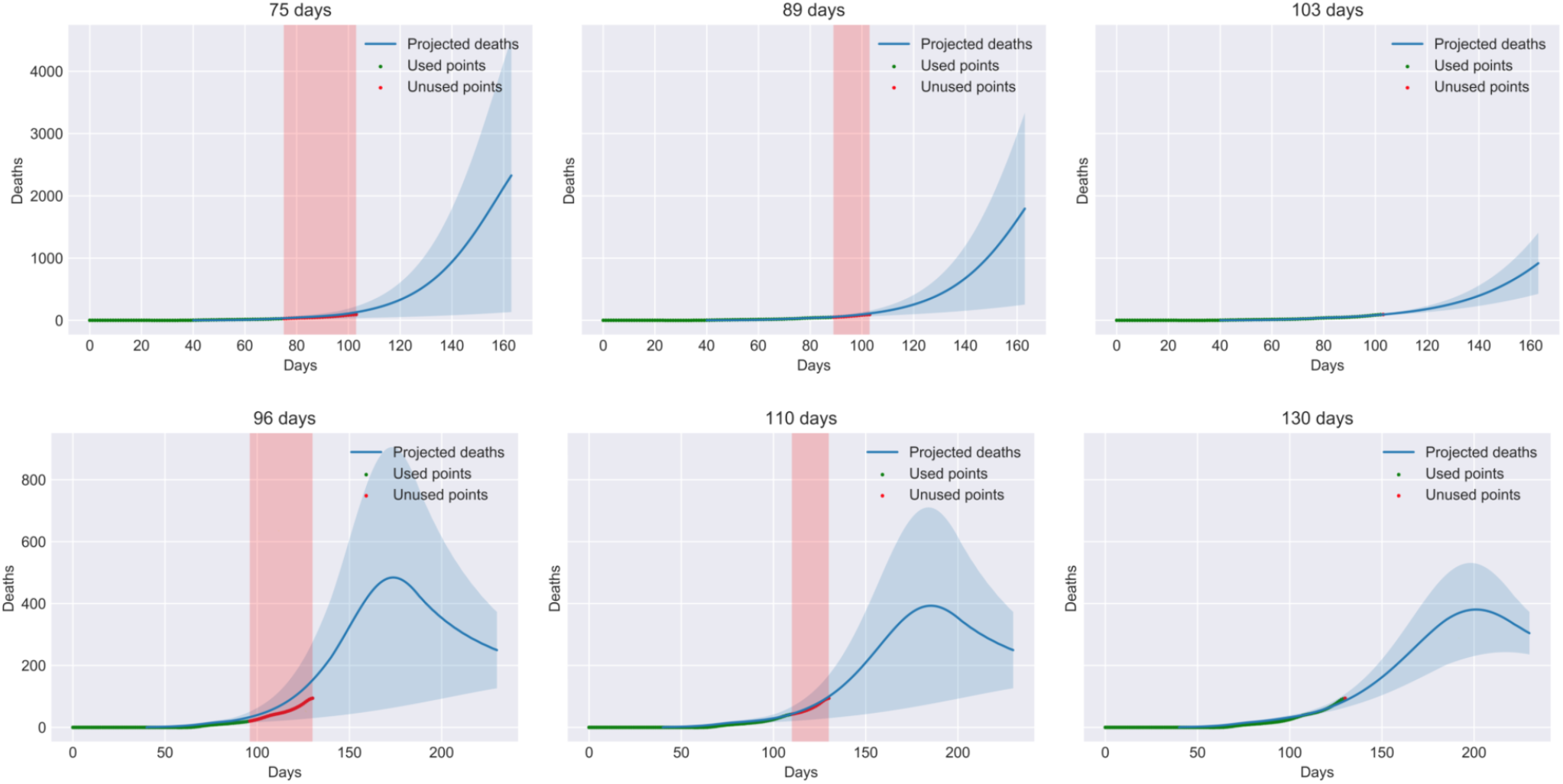
Predictions with uncertainty intervals for Maharashtra and Delhi (from top to bottom). In each figure, the red area contains points the model has not been fitted on and the shaded blue region is a confidence interval. We give projections for the death counts into the future in both cases.

**Table 7:** Model Parameters for Maharashtra

| Breakpoint | Train Loss | Population | $\gamma_X$ | $I_{init}$ | Offset | $R_{max}$ | $R_{min}$ |
| --- | --- | --- | --- | --- | --- | --- | --- |
| 80 | 4.1892 | 114.2 mil | 1.6e-3 | 5e-6 | 40 | 1.6316 | 1.1000 |
| 89 | 3.8369 | 114.2 mil | 1.6e-3 | 5e-6 | 40 | 1.6316 | 1.1105 |
| 103 | 4.0742 | 114.2 mil | 1.6e-3 | 5e-6 | 40 | 1.5737 | 1.2158 |

**Table 8:** Model Parameters for Delhi

| Breakpoint | Train Loss | Population | $\gamma_X$ | $I_{init}$ | Offset | $R_{max}$ | $R_{min}$ |
| --- | --- | --- | --- | --- | --- | --- | --- |
| 102 | 2.7944 | 19 mil | 1.6e-3 | 3.3e-5 | 60 | 1.6894 | 1.3 |
| 106 | 3.5318 | 19 mil | 1.6e-3 | 3.3e-5 | 60 | 1.7473 | 1.3 |
| 110 | 3.8958 | 19 mil | 1.6e-3 | 3.3e-5 | 60 | 1.8052 | 1.3 |

*Rmin* < 1 for Italy, NYC and Spain but *R_min_* > 1 for Maharashtra and Delhi. Now that the country is re-opening *R* can only be expected to increase further. In particular, our model for Delhi predicts that even at lock-down levels of social distancing, the peak is around 75 days out and we can expect to see as many as 400 deaths per day near the peak. This is a horrifying scenario to contemplate and is going to severely affect the elderly, those with co-morbities and our frontline workers.

It is hoped that through these results we are able to emphasize the urgency with which the India needs to find an effective strategy to contain the virus.

### 6.1 Further work

Based on our discussion in the previous sections, we can see the following directions in which the model can be improved

- **Improving quality of** *R* **estimation** With the wide usage of the Aarogya Setu app, the government has accurate raw data available for people’s movement patterns. If we can acquire this data through official channels, we can further improve our *R* estimates.
- **Constructing an online dashboard** We can construct an online dashboard which shows projections with uncertainty intervals in real-time for all districts in India. Further, we can allow the user to transparently adjust *R* to understand how critical social distancing is to contain the spread.
- **Improving uncertainty estimation** Currently we choose *α* based on empirical conditions. Which is to say, we run the model on many different countries and choose *α* for which a large majority of predictions fall within the confidence interval. Can we choose *α* in a more principled manner?
- **Collaboration with MoHFW** A big reason for undertaking this project was that we recognized the urgent need to come up with effective strategies to combat COVID19. Given that IIT Delhi is a well-respected institution we hoped that through the I4 students challenge we would be able to communicate this urgency to the government. If we can collaborate with policy-makers at the Ministry for Health and Welfare we believe this model can help many people in the days to come.

Please feel free to contact the authors or open an issue/pull request on the github repository to request clarification, suggest improvements or features.

## Data Availability

We have used open-source data for death projections from three sources:
John Hopkins CSSE COVID19 repository for daily death count for regions outside India, COVID19India API for daily death count for Indian States and Union Territories, and Community Mobility Data for all regions maintained and made public by Google.

https://github.com/CSSEGISandData/COVID-19

https://www.covid19india.org/

https://www.google.com/covid19/mobility/

